## Supplement 1 (Methods Supplement) for "Impact of the Liberian National Community Health Assistant Program on Childhood Illness Treatment in Grand Bassa County, Liberia: A Difference-in-Differences Analysis of Population-Based Data"

Additional information on sampling and weighting

We used different sampling strategies at baseline and follow-up to simplify the logistics of the substantially larger baseline sample. Both samples were population-based cluster samples which produced population-representative estimates.

The baseline sample was drawn immediately after full household mapping of the county. The sample was stratified by health districts, within which communities were drawn by simple random sample. Within each selected community, we surveyed all households. A drawback to taking a simple random sample of communities in this way is that the total sample size can become unpredictable because communities differ in size, so there is a (relatively small) risk that the final sample will meaningfully vary from the intended sample size. If the sample is meaningfully smaller, the study may be underpowered; if it is larger, the time and funds allocated to the survey may become untenable. We addressed this challenge by drawing communities at random until the selected communities included the cumulative number of households intended for that stratum.

This approach removes the sample size unpredictability but results in larger communities being slightly more likely to be sampled than smaller communities because it is more likely that a larger community will push the cumulative household count over the threshold. However, this problem can be addressed by weighting the data. We estimated the probability a community would be sampled by simulating sampling for each stratum 15,000 times and determining how often each community would be drawn. We then adjusted sampling weights to account for each communities propensity to be over- or under-sampled. The maximum variation in sampling probabilities was 10% in one stratum; in the others it was under 5%. The final sampling weights incorporate both this adjustment and adjustments for different sampling fractions per stratum.

The follow-up survey used a conventional cluster sampling approach identical to that described by White et al. and Ly et al.(1,2) Within each stratum, we selected communities with probability proportional to the number of households in each community. We then selected 24 households in each community. Households were selected with a modified random walk procedure in which a paper triangle was in the center of each community and a house was chosen in the direction of each of the triangle’s points using a random number generator. From there, the next nearest house was chosen until eight households in each segment were selected. Because this design is self-weighting within strata, sampling weights only accounted for different sampling fractions per stratum.

IPTW balance diagnostics.

Though the difference-in-differences framework should account for potential confounders that are either time-invariant or that varied comparably in the intervention and comparison areas, we further adjusted for observable confounders using inverse probability of treatment weighting (IPTW). We did this for two main reasons. First, we could not exclude the possibility of compositional changes over time in the population in ways that differed between intervention and comparison areas, though we did not have reason to expect such changes. Secondly, because we only had one pre-intervention measurement point, we could not assess pre-intervention parallel trends. Adjusting for observable variables reduces the risk bias from different trends in determinants of child health treatment between intervention and comparison areas.

We used two principal approaches for assessing covariate balance in the IPTW models. First, we compared standardized differences (for all variables) and variance ratios (for the continuous variables). All comparisons were against the intervention group at baseline. All standardized differences were less Rubin’s recommendation of absolute values less than 0.25. Similarly, all variance ratios for all continuous variables (distance, the number of household children, child’s age, and maternal age) were between 0.5 and 2.0 as Rubin recommends.(3) Austin and Stuart recommend more stringently that standardized differences have absolute values less than 0.1, which we met by 37 or 39 standardized differences.(4,5) We present these balance diagnostics in more detail in the figure and table below.

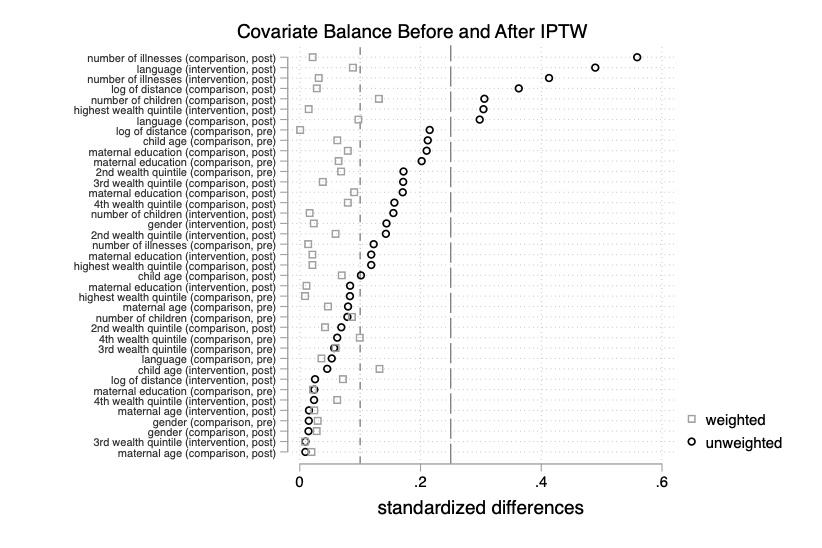

| Table 1. Covariate balance as measured by standardized differences and variance ratios before and after applying inverse probability of treatment weighting. | | | | |
| --- | --- | --- | --- | --- |
|  | Standardized Difference | | Variance Ratio | |
|  | Raw | IPTW | Raw | IPTW |
| **Intervention, post** |  |  |  |  |
| Child gender | -.144 | -.023 |  |  |
| 2nd wealth quintile | .143 | .059 |  |  |
| 3rd wealth quintile | .009 | -.009 |  |  |
| 4th wealth quintile | .024 | -.062 |  |  |
| 5th wealth quintile | -.304 | -.015 |  |  |
| Log of distance | -.025 | -.071 | .646 | .611 |
| Some primary schooling | -.118 | -.021 |  |  |
| Completed primary schooling | -.083 | .011 |  |  |
| Age of child | .045 | .132 | .954 | .947 |
| Age of mother | -.015 | .024 | 1.157 | 1.156 |
| Bassa language | .489 | -.088 |  |  |
| >1 childhood illness | -.413 | -.031 |  |  |
| Number of household children under 5 | -.155 | .016 | .736 | .811 |
| **Comparison, pre** |  |  |  |  |
| Child gender | -.0149 | -.030 |  |  |
| 2nd wealth quintile | .172 | .069 |  |  |
| 3rd wealth quintile | -.057 | .059 |  |  |
| 4th wealth quintile | -.062 | -.099 |  |  |
| 5th wealth quintile | .083 | -.009 |  |  |
| Log of distance | -.215 | -.001 | 1.137 | 1.242 |
| Some primary schooling | .024 | .022 |  |  |
| Completed primary schooling | .202 | .064 |  |  |
| Age of child | .212 | .062 | 1.030 | .976 |
| Age of mother | -.080 | -.047 | 1.042 | 1.031 |
| Bassa language | .053 | -.036 |  |  |
| >1 childhood illness | -.122 | -.014 |  |  |
| Number of household children under 5 | -.079 | .086 | .784 | .920 |
| **Comparison, post** |  |  |  |  |
| Child gender | -.014 | -.028 |  |  |
| 2nd wealth quintile | -.069 | .042 |  |  |
| 3rd wealth quintile | .171 | .038 |  |  |
| 4th wealth quintile | .157 | -.079 |  |  |
| 5th wealth quintile | .118 | .021 |  |  |
| Log of distance | -.363 | -.028 | 1.141 | 1.246 |
| Some primary schooling | -.171 | -.090 |  |  |
| Completed primary schooling | .210 | .079 |  |  |
| Age of child | .101 | .069 | .885 | .910 |
| Age of mother | .009 | -.019 | 1.138 | 1.038 |
| Bassa language | .298 | .097 |  |  |
| >1 childhood illness | -.559 | .021 |  |  |
| Number of household children under 5 | -.306 | .131 | .776 | 1.151 |
| Note: The raw standardized differences and variance ratios incorporate sampling weights. The IPTW values incorporate both sampling weights and inverse probability of treatment weights. All differences are compared to the intervention group prior at baseline. | | | | |

Second, we present an IPT-weighted version of manuscript table 1, which shows the prevalence of each covariate in each time-by-intervention group. This approach allows one to assess the absolute magnitude of any imbalance that remains in covariates after IPT weighting. Because of the difference-in-differences design of our study, the greatest concern would be if there were large residual differences-in-differences in those covariates expected strongly to drive childhood illness treatment after weighting. The largest difference-indifferences for any covariate is 10.2 percentage points for mining community residence, a covariate that could not be included in the propensity score models because of a small number of observations. Even if residence in a mining community completely determined childhood treatment, it could not account for much of the 60 percentage point treatment effect we observed in our main analysis.

| Table 2. Characteristics of Children With Fever, Acute Respiratory Infection, or Diarrhea after IPTW weighting | | | | |
| --- | --- | --- | --- | --- |
|  | Pre-Intervention  (n=888) | | Post-Intervention  (n=395) | |
|  | Intervention (n = 346)  % (95% CI) or Mean  (95% CI) | Comparison  (n = 542)  % (95% CI) or Mean (95% CI) | Intervention (n = 213)  % (95% CI) or Mean (95% CI) | Comparison (n = 182)  % (95% CI) or Mean  (95% CI) |
| Child’s household located in mining community, % | 0 | 12.7  (6.6, 23.0) | 6.9  (1.8, 23.3) | 10.4  (3.6, 26.5) |
| Child's gender, % female | 46.9  (41.0, 52.9) | 45.2  (40.9, 49.7) | 46.7  (40.4, 53.0) | 45.9  (37.5, 54.4) |
| Child’s age in months, mean | 24.5  (22.7, 26.3) | 25.5  (24.3, 26.7) | 26.5  (24.3, 28.8) | 25.8  (23.3, 28.2) |
| Mother’s education , % |  |  |  |  |
| No education | 70.2  (64.2, 75.5 | 66.9  (62.4, 71.1) | 69.2  (60.5, 76.7) | 70.6  (59.9, 79.5) |
| Some primary education | 21.9  (17.9, 26.4) | 23.1  (19.6, 27.0) | 21.8  (15.4, 30.0) | 19.8  (12.4, 30.1) |
| Completed primary or higher | 8.0  (4.6, 13.5) | 10.0  (7.7, 13.0) | 9.0  (4.5, 17.0) | 9.6  (5.7, 15.7) |
| Mother’s survey language, % |  |  |  |  |
| English | 33.4  (22.7, 46.1) | 34.1  (28.6, 40.2) | 34.5  (21.6, 50.2) | 28.8  (17.5, 43.4) |
| Bassa | 66.6  (53.9, 77.3) | 65.9  (59.8, 71.4) | 65.5  (67.6, 87.6) | 71.2  (56.6, 82.5) |
| Mother married or cohabitating, % | 82.9  (77.2, 87.4) | 85.4  (81.7, 88.5) | 92.4  (87.1, 95.6) | 94.7  (89.5, 97.4) |
| Number of illnesses per child in the past 2 wk, % |  |  |  |  |
| 1 illness | 51.5  (45.3, 57.7) | 52.0  (47.1, 56.9) | 52.5  (41.1, 63.6) | 51.9  (43.2, 60.5) |
| ≥ 2 illnesses | 48.5  (42.3, 54.7) | 48.0  (43.1, 52.9) | 47.5  (36.4, 58.9) | 48.1  (39.5, 56.8) |
| Child’s household distance from facility, km, mean | 14.8  (12.5, 17.1) | 15.2  (12.9, 17.5) | 14.2  (12.7, 15.8) | 15.7  (11.5, 19.9) |
| Child’s household wealth index score, mean | -0.02  (-0.40, 0.36) | -0.11  (-0.32, 0.09) | -0.29  (-0.56, -0.01) | -0.09  (-0.47, 0.28) |
| Mother’s no. of children, mean |  |  |  |  |
| Aged < 5 years | 1.4  (1.4, 1.5) | 1.5  (1.4, 1.6) | 1.5  (1.4, 1.6) | 1.5  (1.4, 1.7) |
| Aged < 1 years | 0.4  (0.3, 0.4) | 0.3  (0.3, 0.4) | 0.3  (0.3, 0.4) | 0.3  (0.2, 0.4) |
| Mother’s age in years, mean | 29.0  (28.3, 29.8) | 28.8  (28.1, 29.5) | 28.8  (28.1, 29.5) | 29.2  (27.8, 30.5) |
| Note: The sample sizes in this table are unweighted. The percentages are weighted by both sampling weights and inverse probability of treatment weights. Thus, they should be interpreted as population-representative distributions of each variable after balancing by IPTW. A version of this table showing balance before applying inverse probability of treatment weighting is available in manuscript table 1. | | | | |
