## Supplement 2 (Survey Instrument) for "Impact of the Liberian National Community Health Assistant Program on Childhood Illness Treatment in Grand Bassa County, Liberia: A Difference-in-Differences Analysis of Population-Based Data"

Village name: ______________Today’s date (d/m/y):_______________________

Survey language: __ English __ Kpelle __ Bassa __ French __Other: __________

**ASK THE QUESTIONS IN SECTION 1 TO THE FEMALE HEAD OF HOUSEHOLD; ONLY FILL OUT ONE PER HOUSEHOLD**

**SECTION 1: HOUSEHOLD QUESTIONS**

1.1 ­­­How many persons are living in your house together, with even strangers and visitors who slept here last night? (number ___, this then gives the number of slots for the table below)

| # | First name  [Declined to answer = 888]  [Name is not retained] | Sex | Age  (or birth year)  [Declined to answer = 888] | (NAME) living here all the time? | (NAME) slept here last night? |
| --- | --- | --- | --- | --- | --- |
| 1 |  | - Male - Female - Declined to answer |  | - Yes - No - Declined to answer | - Yes - No - Declined to answer |
| 2 |  | - Male - Female - Declined to answer |  | - Yes - No - Declined to answer | - Yes - No - Declined to answer |
| 3 |  | - Male - Female - Declined to answer |  | - Yes - No - Declined to answer | - Yes - No - Declined to answer |

Any more persons to add? (yes/no)

If yes to above question: How many more persons need to be added? (number ____, will provide that many more slots to fill out)

| 1.2 Is there anyone in this house who get farm land? | - Yes -> 1.3 - No -> 1.4 - Declined to answer -> 1.4 |
| --- | --- |
| 1.3 Number of laps | _________ |
| 1.4 [select one] Where do you people get water from for drinking most times? | - Hand pump, protected well - Tanker truck - Rainwater - public tap/standpipe - Bottled water - Piped into dwelling - Surface water (creek/pond/canal) - Protected spring - tube well or borehole - Piped to yard/plot - Unprotected well - Cart with small tank - Unprotected spring - Other - Declined to answer |
| 1.5 [select one] Where do you people go to toilet/poo poo? | - Pit latrine without slab/open pit - Hanging toilet/hanging latrine - Pit latrine with slab - Flush to pit latrine - Ventilated improved pit latrine - Flush to septic tank - No facility/bush/field - Flush to piped sewer system - Composting toilet - Flush to somewhere else - Bucket toilet - Other - Flush, don’t know where - Declined to answer |
| 1.6 People from other houses go to the same toilet? | - Yes - No - Declined to answer |
| 1.7 [Multiple] Does your household have | - Chairs? - Ice box? - Table? - Electricity? - Cell phone? - Solar panel? - Radio? - Generator? - Cupboard? - Television? - Mattress (not grass)? - Computer? - Sewing machine? - None - Declined to answer |
| 1.8 [select one] What you use for cooking – that coal, gas stove, wood, other thing? | - Wood - Fire coal/charcoal - Gas cylinder - No food cooked in house - Kerosene stove - Biogas - Electricity - Other - Declined to answer |
| 1.9 [Select one] What do you use for light in the house? | - Electricity - Solar - Oil lamp/jackolantern - Gas - Wood - Battery - Kerosene - Chinese lamp - Candles - No light in house - Other - Declined to answer |

| 1.10 [Observe] Please record the main materials of the floor in the house | - Earth/sand/mud - Concrete/Cement - Floormat/Linoleum/vinyl - Ceramic tiles - Wood planks - Carpet - Parquet or polished wood - Other |
| --- | --- |
| 1.11 [Observe] Please record the main materials of the roof of the house | - Thatch/palm leaf - Tarpaulin, plastic - Zinc, metal - Palm/bamboo - Wood planks - Concrete, cement - Wood - Rustic mat - Ceramic tiles - Asbestos sheets/shingles - Other |
| 1.12 [Observe] Please record the main materials of the outside walls of the house | - Cane/palm/trunks - Mud and sticks - Stone blocks - Mud bricks - Bricks - Plywood - Reused wood - Cement - Wood planks/shingles - Straw, thatch mats - Cardboard, plastic - Zinc, metal - Other |
| 1.13 [888=declined to answer] How many bedrooms in this house that people sleep in? | ____ |
| 1.14 [read all choices] [Multiple] Anyone in this house here get: | - A watch - A bicycle - A motorbike - A car/truck - A boat/canoe - None of the above - Declined to answer |
| 1.15 You have any animals, chickens, or ducks in this house? | - Yes -> 1.16 - No -> 1.17 - Declined to answer -> 1.17 |
| 1.16 How many animals people in this house get? | ___ Cows  ___ Pigs  ___Goats  ___ Sheep  ___ Chicken, ducks, or birds |
| 1.17 Anyone in this house who get money in the bank? | - Yes - No - Declined to answer |

Village name: ______________ Name: ____________ Female ID: ____________ Age (or birth year):______Today’s date ___________

Survey language: __ English __ Kpelle __ Bassa __ French __Other __________

**ASK THE QUESTIONS IN SECTION 1 TO EVERY FEMALE AGED 18-49 IN THE HOUSEHOLD**

| 2.1 How long you been living here in this village? | - Yes, always -> 2.2 - Yes, more than 1 year -> 2.1a - Yes, less than 1 year -> 2.1b - Just visiting -> 2.2 - Declined to answer -> 2.2 |
| --- | --- |
| 2.1a If more than 1 year, How many years have you been living in the village? | ________ -> 2.2 |
| 2.1b If less than 1 year, How many months have you been living in the village? | ________ -> 2.2 |
| 2.2 You been to school before? | - Yes -> 2.4 - No -> 2.5a - Declined to answer -> 2.5a |
| 2.4 [888 = declined to answer, 999 = I don’t know] What is the highest grade you completed? | ______ |
| 2.5a You ever born a child? | - Yes -> 2.5b - No, **never had belly** -> 2.26 - Declined to answer -> 2.26 |
| 2.5b From five years up to now, you ever born a child? | - Yes -> 2.6 - No -> 2.26 - Declined to answer -> 2.26 |
| 2.6 When was your last birth? | Mm/dd/yyyy |
| 2.7 Did you see anyone for big belly checkups when you had belly with that child? | - Yes -> 2.8 - No ->2.11 - Declined to answer -> 2.11 |
| 2.8 [Multiple] Who did you see for big belly checkup? Any other person? | - Doctor - Nurse - gCHV - CHW - Physician Assistant (PA) - Certified midwife - Country midwife/Zoe - Trained traditional midwife (TTM) - Other - Declined to answer |
| Other | ______________________________ |
| 2.9 [Multiple] Where did you go to get the checkups? Any other place? | - Home - Other home - Drug store - Clinic/hospital - Other - Declined to answer |
| Other | _________________________________ |
| 2.10 [888 = declined to answer, 999 = don’t know] How many times did you get big belly checkups when you had belly with that child? | _________________________________ |
| 2.11 Where you born the child? | - Clinic/hospital -> 2.12 - Your home -> 2.16 - Other home -> 2.16 - Other -> 2.16 - Declined to answer ->2.19 |
| Other | _________________________________ |
| 2.12 [999 = I don’t know, 888 = declined to answer] How long you stayed in the clinic after you born the child? | Write number: _______  Select unit:   - Weeks - Days - Hours |
| 2.13 After the child was born but before you left the clinic, any doctor or nurse check you to know how you were coming on? | - Yes -> 2.14 - No -> 2.19 - Declined to answer -> 2.19 |
| 2.14 [999 = I don’t know, 888 = declined to answer] How long it took after you born the baby before they first check on you? | Write number: _______  Select unit:   - Weeks - Days - Hours |
| 2.15 [Multiple] Who check on you to see how you were coming on that time? Any other person? | - Clinic staff -> 2.19 - Trained traditional midwife (TTM) -> 2.19 - Country midwife/Zoe -> 2.19 - Other -> other text, then 2.19 - Declined to answer -> 2.19 |
| Other | ________________________________ |
| 2.16 After you born the child, any person check on you to see how you coming on that time? | - Yes -> 2.17 - No -> 2.19 - Declined to answer -> 2.19 |
| 2.17 [999 = don’t know, 888 = declined to answer] How long it took after you born the baby before they first check you? | Write number: _______  Select unit:   - Weeks - Days - Hours |
| 2.18 [Multiple] [Read all choices] Who check on the baby condition at that time? | - Clinic staff - gCHV - TTM - FHW - Country midwife/Zoe - Family/friends - Other - Declined to answer |
| 2.19 The child you born, is it still alive | - Yes -> 2.22 **(less than 2 months -> 2.26)** - No - Declined to answer –> 2.22 **(less than 2 months -> 2.26)** |
| 2.20 When the child die? | - At birth -> 2.26 - After birth -> **2.21** - Declined to answer -> 2.22 |
| 2.21 [999 = I don’t know, 888 = Declined to answer] How many years, months, or days the child get when the child die? | Write number: _______  Select unit:   - Years - Months **(less than 2 months -> 2.26)** - Days |
| 2.22 Any person check on the baby condition in the first two months after the child was born? | - Yes - No -> 2.25b **(less than 2 months -> 2.26)** - I don’t know -> 2.25b **(less than 2 months -> 2.26)** - Declined to answer -> 2.25b **(less than 2 months -> 2.26)** |
| 2.23 [999 = I don’t know, 888 = Declined to answer] How long it look after you born the baby before they first check on his/her condition? | Write number: _______  Select unit:   - Weeks - Days - Hours |
| 2.24 [Multiple] Who check on the baby condition at that time? | - Clinic staff - gCHV - TTM - FHW - Country midwife/Zoe - Family/friends - Other - Declined to answer |
| 2.25 What place they check on the baby first? | - Clinic/hospital - Your home - Other home - Other |
| Other | _____________________________ |
| 2.25b Before your youngest child reached 6 months, that only titi water you give, or you give thing like juice or creek water? | - Baby only had titi water - Baby had other things - Declined to answer |
| 2.26 Now I will like to talk to you about family planning. You marry now-now or you living with a man just like you people marry? | - Yes -> 2.28 - No -> 2.27 - Declined to answer -> 2.27 |
| 2.27 You ever had something to do with a man in this month that passed? | - Yes - No - Declined to answer |
| 2.28 You get belly now-now | - Yes -> 2.29 - No -> 2.30 - I don’t know -> 2.30 - I can’t get pregnant -> 2.43 - Declined to answer -> 2.30 |
| 2.29 This belly you get now, the time it came you wanted it? | - Yes ->2.38 - No ->2.38 - Declined to answer ->2.38 |
| 2.30 You currently doing something or using any method to delay or avoid getting pregnant? | - Yes -> 2.31 - No -> 2.33 - I can’t get pregnant -> 2.43 - Declined to answer -> 2.33 |
| 2.31 [Multiple] Which thing you using? | - Pill - Condom - IUD - Rhythm method - Injection - Withdrawal - Emergency contraception - Other - Declined to answer |
| Other | _______________________________ |
| 2.32 Where did you go the last time for the family planning you taking now? | - Clinic/hospital - gCHV - FHW - Drugstore - Tablet man/black bagger - Country doctor/Zoe - Declined to answer |
| 2.33 You born a child in the last two years? (SKIP if never had a belly) | - Yes -> 2.34 - No -> 2.34 - I can’t get pregnant ->2.43 - Declined to answer ->2.34 |
| 2.34 You now start seeing your time since your last belly? (SKIP if never had a belly) | - Yes - No - Declined to answer |
| 2.35 The last time you get belly, you wanted it that time? (SKIP if never had a belly) | - Yes - No - Declined to answer |
| 2.36 You still want some children? | - Yes - No -> 2.38 - Don’t know -> 2.38 - Can’t get pregnant -> 2.43 - Declined to answer -> 2.38 |
| 2.37 You want to have your next child sometime in the next two years? | - Yes - No - I don’t know - Declined to answer |
| 2.38 You know any place where a person can get some for family planning? | - Yes -> 2.39 - No -> 2.40 - Declined to answer -> 2.40 |
| 2.39 [Multiple] Where is that? | - Drug store - Hospital/clinic - gCHV - FHW - Tablet man/blackbagger - Other - Declined to answer |
| Other | __________________________ |
| 2.40 If you wanted to, could you yourself get something for family planning? | - Yes - No - Don’t know - Declined to answer |
| 2.41 When you last saw your time? | - Never -> 2.43 - Hysterectomy -> 2.43 - Days ago -> 2.42 - Weeks ago -> 2.42 - Months ago -> 2.42 - Years ago -> 2.42 - Declined to answer -> 2.43 |
| 2.42 Time since last menstrual period | Enter number ______  Select unit   - Days - Weeks - Months |
| 2.43 It not that I want to know how it was looking, but you ever to go to do your HIV/AIDS test before? | - Yes - No - Declined to answer |
| 2.44 Have you been seen by a community health worker in the last 3 months | - Yes -> 2.45 - No -> 2.46 - Declined to answer -> 2.46 |
| 2.45 Type of CHW seen | - gCHV - TTM - Country doctor - CHW - Other |
| Other | ______________________________ |
| 2.46 Does X have any children either alive or dead? | - Yes -> 2.47 - Does not have any children -> End of survey - Declined to answer -> End of survey |
| 2.47 How many children does X have either alive or dead? Please include all children you born, even if they died at birth or never get any name. | ______________________________ |

**Birth History**

Now I want you to tell me about all of the children you born, whether still alive or not, starting with the first one. Please include all children you born, even if they died at birth or never get any name.

| What is the first name of your (first/next) child? | Is (NAME) a boy of a girl? | In what month and year was (NAME) born? | Was (NAME) born in a clinic? | Is (NAME) still living? | IF DEAD, how old was (NAME) when he/she died? |
| --- | --- | --- | --- | --- | --- |
|  | - Boy - Girl | Year: _____  Mth: ______ | - Yes - No - Declined to answer | - Yes - No - Declined to answer | Enter number ______  Select unit   - Days - Weeks - Months |
|  | - Boy - Girl | Year: _____  Mth: ______ | - Yes - No - Declined to answer | - Yes - No - Declined to answer | Enter number ______  Select unit   - Days - Weeks - Months |
|  | - Boy - Girl | Year: _____  Mth: ______ | - Yes - No - Declined to answer | - Yes - No - Declined to answer | Enter number ______  Select unit   - Days - Weeks - Months |

Village name: ______________ Mother’s name: ____________ Mother’s Female ID: _____Today’s date (d/m/y):_______________________

**COMPLETE THIS FORM FOR EVERY CHILD UNDER THE AGE OF 5**

Child’s name: _________________________ Child’s gender:________________________

| 3.1 Child’s date of birth | ___ ___ / ___ ___ / ___ ___ ___ ___ |
| --- | --- |
| 3.2 [NAME] ever had running stomach in these two weeks that passed? | - Yes -> 3.3 - No -> 3.9 - Don’t know -> 3.9 - Declined to answer -> 3.9 |
| 3.3 You got treatment from anywhere or anyone the time [NAME] stomach was running? | - Yes -> 3.4 - No -> 3.7 - Declined to answer -> 3.7 |
| 3.4 [Multiple] Where or who [NAME] got treatment from? Any other place? | - Drug store - Tablet man/blackbagger - gCHV - Hospital/clinic - Country doctor - Zoe - CHW - Other - Declined to answer |
| Other | __________________________________ |
| 3.5 Who did you go for treatment FIRST? | - Drug store - Tablet man/blackbagger - gCHV - Hospital/clinic - Country doctor - Zoe - CHW - Other - Declined to answer |
| Other | ____________________________ |
| 3.6 [999 = don’t know, 888 = declined to answer] When the stomach started running, how many days it took before [NAME] got treatment? | ____________ |
| 3.7 Since [NAME]’s stomach started running, anybody give him/her glucose water (ORS) to drink? | - Yes - No - I don’t know - Declined to answer |
| 3.8 Since [NAME]’s stomach started running, anybody give him/her homemade sugar/salt drink? | - Yes - No - I don’t know - Declined to answer |
| 3.9 [NAME] ever get hot (fever) in these two weeks that passed? | - Yes -> 3.10 - No -> 3.12 - I don’t know ->3.12 - Declined to answer ->3.12 |
| 3.10 When [NAME]’s skin was hot, anyone take blood from his/her finger or hell to do malaria test? | - Yes -> 3.11 - No -> 3.12 - I don’t know -> 3.12 - Declined to answer -> 3.12 |
| 3.11 [NAME]’s test showed that he/she got malaria? | - Yes - No - I don’t know - Declined to answer |
| 3.12 [NAME] ever got cough in these two weeks that passed? | - Yes -> 3.13 - No -> 3.15 if had fever, 3.20 if no fever - I don’t know -> 3.15 if had fever, 3.20 if no fever - Declined to answer -> 3.15 if had fever, 3.20 if no fever |
| 3.13 That time [NAME] was coughing, you saw him/her breathing fast-fast or he/she was catching hard time to breathe? | - Yes -> 3.14 - No -> 3.15 - I don’t know -> 3.15 - Declined to answer -> 3.15 |
| 3.14 You think the fast breathing was caused by some problem in [NAME]’s chest or something closing [NAME]’s nose or both? | - Chest only - Nose only - Both chest and nose - I don’t know - Other - Declined to answer |
| 3.15 You got treatment from anywhere or anyone the time [NAME] was sick? | - Yes -> 3.16 - No -> 3.20 - Declined to answer -> 3.20 |
| 3.16 [Multiple] Where or who [NAME] got treatment from? Any other place? | - Drug store - Tablet man/blackbagger - gCHV - Hospital/clinic - Country doctor - Zoe - CHW - Other - Declined to answer |
| Other | __________________________ |
| 3.17 Who did you go for treatment FIRST? | - Drug store - Tablet man/blackbagger - gCHV - Hospital/clinic - Country doctor - Zoe - CHW - Other - Declined to answer |
| 3.18 [999 = don’t know, 888 = declined to answer] When the sickness started, how many days it took before [NAME] get treatment? | ______ |
| 3.19 [Multiple] What kind of medicine [NAME] took? Any other medicine? | - New malaria tablet/ACT - Chloroquine - Country medicine - Antibiotics - Don’t know - Other - Declined to answer |
| Other | _____________________________ |
| 3.20 [NAME] sleep under mosquito net last night? | - Yes - No - I don’t know - Declined to answer |
| 3.21 You get vaccine card for [NAME] | - Yes -> 3.22 - No -> 3.26 - Declined to answer -> 3.26 |
| 3.22 I can please see it? | - Yes they gave me the card -> 3.23 - No -> 3.26 |
| 3.23 [Multiple] Which vaccines are marked on [NAME]’s vaccine card? | - BCG -> 3.28 - Penta 1 -> 3.29 - Penta 2 -> 3.30 - Penta 3 -> 3.31 - Polio (P0) -> 3.24 - Polio (P1) -> 3.25 - Polio (P2) -> 3.26 - Polio (P3) -> 3.27 - Measles -> 3.32 - Yellow fever -> 3.33 - Rota 1 -> 3.34 - Rota 2 -> 3.35 - Rota 3 ->3.36 - Pneumo 1 -> 3.37 - Pneumo 2 -> 3.38 - Pneumo 3 -> 3.39 - IPV 1 ->3.40 - IPV 2 ->3.41 - IPV 3 ->3.42   *Go to appropriate data questions for ALL vaccines reported, then go to question 3.43* |
| Date of OPV 0 vaccine on card | ___ ___ / ___ ___ / ___ ___ ___ ___ |
| Date of OPV 1 vaccine on card | ___ ___ / ___ ___ / ___ ___ ___ ___ |
| Date of OPV 2 vaccine on card | ___ ___ / ___ ___ / ___ ___ ___ ___ |
| Date of OPV 3 vaccine on card | ___ ___ / ___ ___ / ___ ___ ___ ___ |
| Date of BCG vaccine on card | ___ ___ / ___ ___ / ___ ___ ___ ___ |
| Date of PENTA 1 vaccine on card | ___ ___ / ___ ___ / ___ ___ ___ ___ |
| Date of PENTA 2 vaccine on card | ___ ___ / ___ ___ / ___ ___ ___ ___ |
| Date of PENTA 3 vaccine on card | ___ ___ / ___ ___ / ___ ___ ___ ___ |
| Date of MEASLES vaccine on card | ___ ___ / ___ ___ / ___ ___ ___ ___ |
| Date of YELLOW FEVER vaccine on card | ___ ___ / ___ ___ / ___ ___ ___ ___ |
| Date of ROTA 1 vaccine on card | ___ ___ / ___ ___ / ___ ___ ___ ___ |
| Date of ROTA 2 vaccine on card | ___ ___ / ___ ___ / ___ ___ ___ ___ |
| Date of ROTA 3 vaccine on card | ___ ___ / ___ ___ / ___ ___ ___ ___ |
| Date of PNEUMO 1 vaccine on card | ___ ___ / ___ ___ / ___ ___ ___ ___ |
| Date of PNEUMO 2 vaccine on card | ___ ___ / ___ ___ / ___ ___ ___ ___ |
| Date of PNEUMO 3 vaccine on card | ___ ___ / ___ ___ / ___ ___ ___ ___ |
| Date of IPV 1 vaccine on card | ___ ___ / ___ ___ / ___ ___ ___ ___ |
| Date of IPV 2 vaccine on card | ___ ___ / ___ ___ / ___ ___ ___ ___ |
| Date of IPV 3 vaccine on card | ___ ___ / ___ ___ / ___ ___ ___ ___ |
| 3.24 [NAME] ever take any other vaccine that they didn’t write on the vaccine card? Like those ones they can give all over the country? | - Yes -> 3.25 - No -> End survey - Don’t know -> End survey - Declined to answer -> End survey |

| 3.25 [Multiple] Which ones? | - BCG - Penta 1 - Penta 2 - Penta 3 - Polio (P0) - Polio (P1) - Polio (P2) - Polio (P3) - Measles - Yellow fever - Rota 1 - Rota 2 - Rota 3 - Pneumo 1 - Pneumo 2 - Pneumo 3 - IPV 1 - IPV 2 - IPV 3 - Don’t know - Declined to answer - Go to end of survey |
| --- | --- |
| 3.26 [NAME] ever take any vaccine to prevent him/her from getting diseases, at the clinic or like those ones they can give all over the country? | - Yes -> 3.27 - No -> End survey - Don’t know -> End survey - Declined to answer -> End survey |
| 3.27 [NAME] ever take the TB vaccine that can make a mark on the child’s arm or shoulder? | - Yes - No - Don’t know - Declined to answer |
| 3.28 [NAME] ever take the vaccine for the sickness that can make children cripple they call polio? That the vaccine they can drop in the child’s mouth. | - Yes -> 3.29 - No -> 3.31 - Don’t know -> 3.31 - Declined to answer -> 3.31 |
| 3.29 How old was [NAME] when they give him/her the first vaccine in the mouth? It was in the first two weeks after he/she was born or it was more than two weeks? | - First two weeks - Later - I don’t know - Declined to answer |
| 3.30 [999 = don’t know, 888 = declined to answer] How many times they now put the polio vaccine in [NAME]’s mouth since he/she was born? | ______ |
| 3.31 They ever give [NAME] the pentavalent vaccine injection on the thigh? They sometimes give it the same time they can put the polio vaccine in the mouth. | - Yes -> 3.32 - No -> 3.33 - I don’t know -> 3.33 - Declined to answer -> 3.33 |
| 3.32 [999 = don’t know, 888 = declined to answer] How many times they give [NAME] pentavalent vaccine injection on the thigh? | ______ |
| 3.33 They ever give [NAME] an injection in the thigh to prevent pneumonia? | - Yes -> 3.34 - No -> 3.35 - I don’t know -> 3.35 - Declined to answer -> 3.35 |
| 3.34 [999 = don’t know, 888 = declined to answer] How many times they give [NAME] injection in the thigh to prevent pneumonia? | _______ |

| 3.35 They ever give [NAME] a rotavirus vaccination, that is, a liquid in the mouth to prevent diarrhea? | - Yes -> 3.36 - No -> 3.37 - I don’t know -> 3.37 - Declined to answer -> 3.37 |
| --- | --- |
| 3.36 [999 = don’t know, 888 = declined to answer] How many times did [NAME] get the rotavirus vaccine? | _________ |
| 3.37 They ever give [NAME] vaccine for measles sickness? That the vaccine injection they can give the children in the arm when they are 9 months or more | - Yes - No - I don’t know - Declined to answer |
| 3.38 They ever give [NAME] vaccine to prevent yellow fever? That the vaccine injection they can give the children in the arm when they are 9 months or more. They sometimes give it the same time they can give the measles vaccine. | - Yes - No - I don’t know - Declined to answer |

Village name: ______________ Woman’s name: ____________ Female ID: ___________Today’s date (d/m/y):_______________________

**COMPLETE THIS QUESTIONNAIRE ONCE PER HOUSEHOLD**

| 4.1 [999 = don’t know, 888 = declined to answer] In the last four weeks, how many times have you or anyone in this household visited or used the following: | gCHV ______  CHW ______  Hospital/Clinic _____  Drugstore _____  Tablet man/black bagger _____  Country doctor _____ |
| --- | --- |
| 4.2 Have you visited any other person or place for your health? | - Yes - No - Declined to answer |
| Other place visited for health | ________________________________ |
| [999 = don’t know, 888 = declined to answer] In the last four weeks, how many times have you or anyone in this household visited or used [OTHER]? | ___________ |
| 4.3 [Multiple] Where do you go for medical advice or treatment? | - To see doctor/nurse at clinic/hospital - To the drugstore - Tablet man/black bagger - gCHV - CHW - Country doctor - Other - Don’t know - Declined to answer |
| Other | _____________________________ |
| 4.4 In the last year, you or anyone in your family ever need medical advice but not able to get it? | - Yes -> 4.5 - No -> 4.6 - Declined to answer -> 4.6 |
| 4.5 [Don’t read list] [Multiple] What things stopping you from getting medical treatment? Any other thing? | - No transport money - No money for treatment - Afraid to go to provider - No place to stay near provider - Distance to provider - No medicine available - No permission to go - No one to take care of family - Provider not available - Ebola outbreak - Other - Declined to answer |
| Other | ______________________________ |
| 4.6 [Do not read list] [Multiple] In your own thinking, what are the signs of someone who can have ebola? | - Fever - Muscle pains - Vomiting - Sore throat - Running stomach - Bleeding from eyes, mouth, nose - Ebola is not real - Other - Don’t know - Declined to answer |
| Other | ___________________________________ |
| 4.7 Can people get ebola from touching an ebola patient? | - Yes - No - Don’t know - Declined to answer |
| 4.8 Can people get ebola from the air? | - Yes - No - Don’t know - Declined to answer |
| 4.9 Can people get ebola by touching or washing a dead body? | - Yes - No - Don’t know - Declined to answer |
| 4.10 Can people get ebola from the vomit of an ebola patient? | - Yes - No - Don’t know - Declined to answer |
| 4.11 Can people get ebola from witchcraft or something like that? | - Yes - No - Don’t know - Declined to answer |
| 4.12 Can people get ebola by going to the hospital/clinic? | - Yes - No - Don’t know - Declined to answer |
| 4.13 If it looking like you or someone in your family have ebola, would you try to get medical advice or treatment? | - Yes -> 4.14 - No -> End survey - Don’t know -> End survey - Declined to answer -> End survey |
| 4.14 [Read all] Where would you go first? | - To see family member, friend, or neighbor - To see CHW - To see gCHV - To see doctor or nurse at the clinic - To the ETU - To the tablet man/black bagger - Country doctor - Other - Don’t know - Declined to answer |
| Other | __________________________ |
