## Supplement 3 (Sensitivity Analyses) for "Impact of the Liberian National Community Health Assistant Program on Childhood Illness Treatment in Grand Bassa County, Liberia: A Difference-in-Differences Analysis of Population-Based Data"

Regression adjustment: This analysis replicates the main analysis except that confounders are adjusted by regression rather than inverse probability of treatment weighting.

Table 1. Difference-in-Differences in Childhood Illness Treatment

|  | Sample size  (unweighted) | | Unadjusted Model | | Regression-Adjusted Model | |
| --- | --- | --- | --- | --- | --- | --- |
| Care-seeking from qualified provider | Pre | Post | DID %  (95% CI) | P | DID %  (95% CI) | P |
| Any illness | 894 | 397 | 55.2  (41.2, 69.3) | <0.001 | 56.8  (42.9, 70.7) | <0.001 |
| Fever | 691 | 293 | 60.7  (45.1, 76.2) | <0.001 | 64.5  (49.6, 79.3) | <0.001 |
| Diarrhea | 590 | 215 | 69.8  (54.5, 85.1) | <0.001 | 67.2  (52.2, 82.2) | <0.001 |
| Acute Respiratory Illness | 295 | 79 | 39.1  (10.9, 67.2) | 0.007 | 38.5  (13.6, 63.4) | 0.003 |
| Oral rehydration therapy for diarrhea | 586 | 215 | 32.9  (15.0, 50.7) | <0.001 | 32.1  (14.8, 49.4) | <0.001 |
| Rapid diagnostic test for fever | 686 | 293 | 41.9  (26.0, 57.8) | <0.001 | 43.5  (28.3, 58.6) | <0.001 |

Restricted only to agricultural areas: This analysis replicates the main analysis except that it excludes respondents who live in mining communities. The main analysis could not adjust for community type by IPTW because too few respondents were from mining communities.

Table 2. Difference-in-Differences in Childhood Illness Treatment

|  | Sample size  (unweighted) | | Unadjusted Model | | Inverse Probability of Treatment Weighted Model | |
| --- | --- | --- | --- | --- | --- | --- |
| Care-seeking from qualified provider | Pre | Post | DID %  (95% CI) | P | DID %  (95% CI) | P |
| Any illness | 825 | 360 | 53.2  (38.7, 67.7) | <0.001 | 61.0  (45.2, 76.9) | <0.001 |
| Fever | 636 | 264 | 58.5  (42.2, 74.8) | <0.001 | 64.9  (47.5, 82.3) | <0.001 |
| Diarrhea | 547 | 199 | 69.5  (54.7, 84.4) | <0.001 | 75.5  (60.0, 90.9) | <0.001 |
| Acute Respiratory Illness | 275 | 73 | 37.6  (7.9, 67.2) | 0.013 | 50.1  (18.3, 81.9) | 0.002 |
| Oral rehydration therapy for diarrhea | 543 | 199 | 30.7  (12.4, 48.9) | 0.001 | 37.5  (19.0, 56.1) | <0.001 |
| Rapid diagnostic test for fever | 631 | 264 | 39.4  (22.6, 56.1) | <0.001 | 38.2  (18.8, 57.5) | <0.001 |

Does not exclude one enumerator’s data: In the main analysis, we excluded data from one enumerator because of data quality concerns. This analysis replicates the main analysis except that it includes that enumerator’s data.

Table 3. Difference-in-Differences in Childhood Illness Treatment

|  | Sample size  (unweighted) | | Unadjusted Model | | Inverse Probability of Treatment Weighted Model | |
| --- | --- | --- | --- | --- | --- | --- |
| Care-seeking from qualified provider | Pre | Post | DID %  (95% CI) | P | DID %  (95% CI) | P |
| Any illness | 894 | 463 | 55.4  (42.5, 68.2) | <0.001 | 59.4  (45.6, 73.2) | <0.001 |
| Fever | 691 | 351 | 60.4  (46.1, 74.7) | <0.001 | 64.7  (50.1, 79.3) | <0.001 |
| Diarrhea | 590 | 277 | 65.4  (51.4, 79.4) | <0.001 | 68.0  (52.5, 83.4) | <0.001 |
| Acute Respiratory Illness | 295 | 127 | 50.4  (28.5, 72.2) | <0.001 | 55.4  (32.5, 78.4) | <0.001 |
| Oral rehydration therapy for diarrhea | 586 | 277 | 40.3  (24.8, 55.8) | <0.001 | 39.5  (23.4, 55.6) | <0.001 |
| Rapid diagnostic test for fever | 686 | 350 | 39.8  (24.1, 55.5) | <0.001 | 37.2  (19.4, 55.0) | <0.001 |
